# Validation of the Cognitive Assessment Instrument for Obsessions and Compulsions (CAIOC-13) in an Indian Sample

**DOI:** 10.1101/2023.06.12.23291270

**Authors:** Mahashweta Bhattacharya, Himani Kashyap, Srinivas Balachander, YC Janardhan Reddy

**Affiliations:** Obsessive-Compulsive Disorder Clinic, Department of Psychiatry, National Institute of Mental Health and Neurosciences (NIMHANS), Bengaluru, India; Department of Clinical Psychology, National Institute of Mental health and Neurosciences (NIMHANS), Bengaluru, Karnataka, India; Department of Psychiatry, National Institute of Mental Health and Neurosciences (NIMHANS), Bengaluru, Karnataka, India; Accelerator Program for Discovery in Brain Disorders using Stem cells (ADBS), Department of Biotechnology (DBT), Government of India

**Keywords:** Obsessive compulsive disorder, Cognitive assessment, scale validation, self-report measures of cognition

## Abstract

**Background:** Brief self-report measures of cognition are advantageous for flagging significant cognitive dysfunction and minimising the need for extensive neuropsychological assessments. The Cognitive Assessment Instrument for Obsessions and Compulsions (CAIOC-13) is a recently developed 13-item self-rated scale, assessing everyday functional difficulties resulting from cognitive dysfunction specific to those reported by individuals with Obsessive-Compulsive Disorder (OCD) e.g., difficulties with reading, making choices, slowness, perfectionism & procrastination). This study was undertaken to validate the CAIOC-13 in an Indian sample of individuals with OCD.

**Material and Methods:** A total of 75 subjects with OCD and 81 non-clinical controls completed the CAIOC-13, the Perceived Deficits Questionnaire (PDQ) and the Dysfunctional Attitude Scale-Short Form (DAS-SF1). Pearson’s r correlation was used to establish the convergent and divergent validity with PDQ and DAS-SF1 respectively; the Receiver Operating Characteristic (ROC) curve was used to analyze the discriminant validity, and the factorial structure was evaluated using the Principal Component Analysis (PCA).

**Results:** CAIOC-13 scale scores showed a strong significant correlation (r = 0.56 p <0.001) with PDQ scores and a weak correlation with DAS-SF1 scores (r = 0.33 p = 0.003). Area Under the ROC curve (AUC) value was found to be 0.92 indicating that the CAIOC-13 could accurately discriminate between OCD and controls. The PCA analysis also showed a strong loading on a single component.

**Conclusion:** The results suggest that the CAIOC-13 is a valid tool for briefly assessing cognitive deficits in individuals with OCD in India. Future studies may also examine the correlation of CAIOC-13 with standardized neuropsychological assessments.

## Introduction

Cognitive deficits are well established in obsessive-compulsive disorder (OCD), (1) and may precede onset of OCD symptomatology, persist despite remission (2,3) and even predict poor treatment outcome (4). Mounting evidence in other disorders points to cognitive dysfunction as a major contributor to poor clinical and functional outcomes, even when severity of illness is controlled for (5). OCD is associated with significant academic and occupational dysfunction (6), and treatments targeting cognition may enhance occupational functioning in OCD (7), however the relationship between cognitive and occupational dysfunction has received scant research attention. Cognitive difficulties in OCD are not routinely assessed in clinical settings, perhaps owing to the time- and labour-intensive nature of neuropsychological assessments. Indeed, despite the objectivity and rigorous psychometric properties of neuropsychological tests, test performance may not always closely correspond to real-life functioning (8). Moreover, research in OCD highlights significant heterogeneity and inconsistency in findings across studies (9) and a recent review has suggested that methodological and psychometric issues inherent to neuropsychological testing such as utilisation of different tests under the same neuropsychological domain, choice of outcome measures, etc may contribute to such heterogeneity (1). In addition to this, discrepancies between objectively assessed neuropsychological performance and subjectively reported cognitive problems are well-known in many disorders (10,11). This indicates the critical need for brief and valid assessments of cognition from multiple perspectives, including self-rating, standardised neuropsychological assessment, and functional correlates. Existing scales to assess subjective report of cognitive difficulties (12,13) are not specific to the difficulties evinced by individuals with OCD.

In this context, the Cognitive Assessment Instrument of Obsessions and Compulsions (CAIOC-13) (14) is a measure developed to assess everyday functional difficulties resulting from cognitive dysfunction specific to OCD (e.g., difficulties with reading, making choices, slowness, perfectionism, procrastination, etc), and has demonstrated good test-retest reliability and discriminant validity in discriminating between OCD, depression, and healthy controls (15).

The CAIOC-13 is likely to be useful for a brief assessment of cognitive functions in clinical settings. The aim of the present study is to validate the CAIOC-13 on an Indian sample of patients with OCD.

## METHOD

### Study setting and design

The study was carried out at the OCD Clinic of a tertiary psychiatric institute in India between January to November 2021 and was approved by the institute ethics committee. A cross-sectional design was adopted and informed consent was obtained from all participants prior to participation.

### Participants

Using power analysis (G-power) (16), the number of participants needed to achieve a significance level of 0.05 with 90% power using a medium effect size for an independent sample t test comparing OCD and controls was estimated at 70 per group. Accordingly, 150 individuals (75 patients diagnosed with OCD according to DSM-5 and 75 non-clinical controls (scoring less than 20 as per the Kessler Psychological Distress Scale K10) aged 18 to 50 years with a working knowledge of English language were recruited for the study. For the OCD group, individuals were excluded if they had a history of other conditions that may exaggerate cognitive impairment, such as bipolar disorder, schizophrenia, substance dependence, central nervous system conditions (Cerebrovascular accident, traumatic brain injury, epilepsy, degenerative disorders), or developmental disorders (intellectual development or specific learning disability, Autism, Asperger’s syndrome, and attention deficit hyperactivity disorder). The OCD sample was recruited from individuals seeking treatment at the tertiary centre. Non-clinical controls were recruited through a flyer shared via email or social media post.

### Materials

- Yale Brown Obsessive Compulsive Scale (Y-BOCS) (17) is a 10-item clinician rated measure of severity of OCD, with a maximum score of 40 (maximum 20 each on obsessions and compulsions).
- Hamilton Anxiety Rating Scale (HAM-A) (18) consists of 14 clinician rated questions to provide a measure of global anxiety that is weighted towards somatic and autonomic features of anxiety, but also includes emotional and cognitive symptoms.
- Montgomery-Asberg Depression Rating Scale (19) consists of 10 items evaluating core symptoms of depression. Nine of the items are based upon patient report, and one is on the clinician’s observation during the rating interview. MADRS items are rated on a 0–6 continuum (0=no abnormality, 6=severe).
- Kessler Psychological Distress Scale (K-10) (20), involves 10 questions about emotional states each with a five-level response scale (one ‘none of the time’ to five ‘all of the time’), yielding a minimum score of 10 and a maximum score of 50. A score above 24 has been suggested as optimum to indicate presence of a psychiatric disorder (21).
- Cognitive Assessment Instrument of Obsessions and Compulsions (CAIOC-13) (14) is a 13-item self-rated dimensional measure of cognitive symptoms in individuals with OCD. It includes functional anomalies, such as difficulties with reading, slowness, doubts, perfectionism and making choices. Responses can be rated from 0 (no difficulty) to 6 (extreme difficulty), providing a maximum total of 78, and a suggested cut-off of 38.5 indicating significant impairments (sensitivity: 70.0%; specificity: 71.7%). The CAIOC-13 has shown good test-retest reliability (ranging from .50 to .91 with 10 items >0.7, total scores 0.91).
- Perceived Deficits questionnaire (PDQ) (22) is a 20-item questionnaire developed to measure the degree to which people perceive themselves to be experiencing cognitive difficulties, in 4 areas - attention/concentration, retrospective memory, prospective memory, and organization/ planning. As there is no existing measure for self-reported cognitive difficulties in OCD, this scale was chosen as the most similar measure for convergent validity.
- Dysfunctional Attitude Scale – Short Form 1 (DAS-SF1; (23) is a 9-item scale to elicit dysfunctional cognitions, subtyped into categories of Dependency, Self-control and Achievement. As dysfunctional cognitions are expected to be dissimilar to self-reported cognitive difficulties, but still within the broad domain of cognition, this scale was chosen for divergent validity.

### Procedure

Given the covid-19 pandemic, and based on individual preferences, the scales were either administered in-person or through an online link. Among the mentioned scales YBOCS, HAM A, MADRS was used only on the OCD group and K-10 scale was used only for the non-clinical controls. Individuals in the control group scoring more than 20 on K-10 were excluded from the analysis. All the other scales were administered on both the groups.

### Data Analysis

The data was analysed using R version 4.2.0. Descriptive statistics of clinical and demographic variables such as means, standard deviations, frequency and percentages was calculated. Validity of the test was established with the following methods - convergent validity through correlation with PDQ; divergent validity through correlation with DAS-SF1; discriminant validity through significance of difference between clinical and control samples using a t-test and the receiver-operating characteristic (ROC) curve. The area under the ROC curve (AUC) was computed as a measure of how accurately the CAIOC-13 could discriminate between OCD and non-clinical controls. The sensitivity and specificity at the different cut-scores were examined to arrive at the optimal cut-off value at which the instrument discriminates cases and control. Factor analysis (principal components analysis) was used to establish factorial validity.

## RESULTS

### Sociodemographic and clinical characteristics

Of the 81 responses received from the non-clinical group, 11 individuals scored greater than 20 on the K10 scale and hence their data were excluded from any further analysis.

The OCD and non-clinical groups were comparable on age, gender, and demographic characteristics except for significantly higher education in the non-clinical group (Table 1). The OCD group had a range of illness severity between moderate to severe. Since the mean anxiety and depression scores of the OCD group lay in the clinically significant range, HAM-A and MADRS scores were considered covariates for further analysis. Lifetime diagnosis of major depressive disorder (12%) and any anxiety disorder (8%) were the most common comorbid illness in the OCD group.

**Table 1:** Demographic variables of the groups:

| Variable | OCD Group<br>(n=75) | NC Group<br>(n=81) | t /chi-square<br>value | p value |
| --- | --- | --- | --- | --- |
| Age [Mean (SD)] | 32.28 (9.30) | 31.88 (7.02) | 0.307 | 0.759 |
| Sex (Male%) | 50 (66.7) | 43 (53.1) | 2.445 | 0.118 |
| Years of Education<br>[Mean (SD)] | 14.83 (2.40) | 17.57 (2.12) | -7.570 | **<0.001** |
| Residence (Urban%) | 62 (82.7) | 60 (74.1) | 1.2204 | 0.269 |
| OCD: Obsessive compulsive Disorder, NC: Non- Clinical Control |  |  |  |  |

**Table 2:** Clinical measures for the two groups:

| Variable | OCD (n=75) | NC (n=81) | t value | p value |
| --- | --- | --- | --- | --- |
| YBOCS Total [mean (SD)] | 22.85 (6.49) | - | - | - |
| HAM-A Total [mean (SD)] | 11.05 (5.81) | - | - | - |
| MADRS Total [mean (SD)] | 16.55 (8.38) | - | - | - |
| K10 Total [mean (SD)] | - | 16.22 (6.55) | - | - |
| % scoring >20 | - | 11 | - | - |
| CAIOC Total [mean (SD)] | 42.16 (12.50) | 11.74 (13.92) | 13.976 | **<0.001** |
| PDQ Total [(mean (SD)] | 32.08 (14.84) | 12.84 (11.93) | 8.851 | **<0.001** |
| DAS-SF1 Total [(mean (SD)] | 20.84 (3.70) | 17.84 (6.78) | 3.386 | **0.001** |
| OCD: Obsessive compulsive Disorder, NC: Non- Clinical Control, YBOCS: Yale Brown Obsessive Compulsive Scale, HAM-A: Hamilton Anxiety Rating Scale, MADRS: Montgomery-Asberg Depression Rating Scale, K10: Kessler Psychological Distress Scale, CAIOC-13: Cognitive Assessment Instrument of Obsessions and Compulsions, PDQ: Perceived Deficits questionnaire, DAS-SF1: Dysfunctional Attitude Scale – Short Form 1 |  |  |  |  |

**Table 3:** Pearson’s correlations between all the scales.

| Variable | YBOCS<br>Total | HAM-A<br>Total | MADRS<br>Total | CAIOC-13 Total | PDQ<br>Total | DAS-SF1<br>Total |
| --- | --- | --- | --- | --- | --- | --- |
| **YBOCS**<br>**Total** | 1(NA) |  |  |  |  |  |
| **HAM-A**<br>**Total** | 0.241*<br>(0.036) | 1(NA) |  |  |  |  |
| **MADRS**<br>**Total** | 0.144<br>(0.213) | 0.628***<br>(<0.001) | 1(NA) |  |  |  |
| **CAIOC-**<br>**13 Total** | 0.110<br>(0.345) | 0.237*<br>(0.040) | 0.404***<br>(< 0.001) | 1(NA) |  |  |
| **PDQ**<br>**Total** | 0.067<br>(0.566) | 0.325**<br>(0.004) | 0.325**<br>(0.004) | 0.526***<br>(< 0.001) | 1(NA) |  |
| **DAS-SF1**<br>**Total** | -0.004<br>(0.975) | 0.290*<br>(0.011) | 0.290*<br>(0.011) | 0.339***<br>(< 0.001) | 0.430**<br>(<0.001) | 1(NA) |
| * p < .05, ** p < .01, *** p < .001<br>YBOCS: Yale Brown Obsessive Compulsive Scale, HAM-A: Hamilton Anxiety Rating Scale, MADRS: Montgomery-Asberg Depression Rating Scale, CAIOC-13: Cognitive Assessment Instrument of Obsessions and Compulsions, PDQ: Perceived Deficits questionnaire, DAS-SF1: Dysfunctional Attitude Scale – Short Form 1 |  |  |  |  |  |  |

### Convergent Validity

Using Pearson’s correlational analysis, the total scores on CAIOC-13 in the OCD group correlated strongly with the total scores on PDQ (0.526, p<0.001).

### Divergent Validity

Using Pearson’s correlational analysis, the total scores on CAIOC-13 in the OCD group showed a low to moderate correlation with the total scores on the DAS-SF1 (0.339), but the correlation was significant with p value= 0.003.

### Discriminant Validity

The ROC analysis gave a cut-score of 28.5 which indicates the optimal combination of sensitivity (accurately identifying true case positives: OCD) and specificity (accurately identifying true case negatives: non-clinical controls). AUC value was found to be 0.92 (An AUC value of 0.50 reflects identification of cases at chance level and 1.0 indicates a perfect diagnostic tool). Even after adjustment for years of education between the two groups, the AUC value was found to be 0.85 (95% CI, 0.798-0.920).

**Figure 1:**
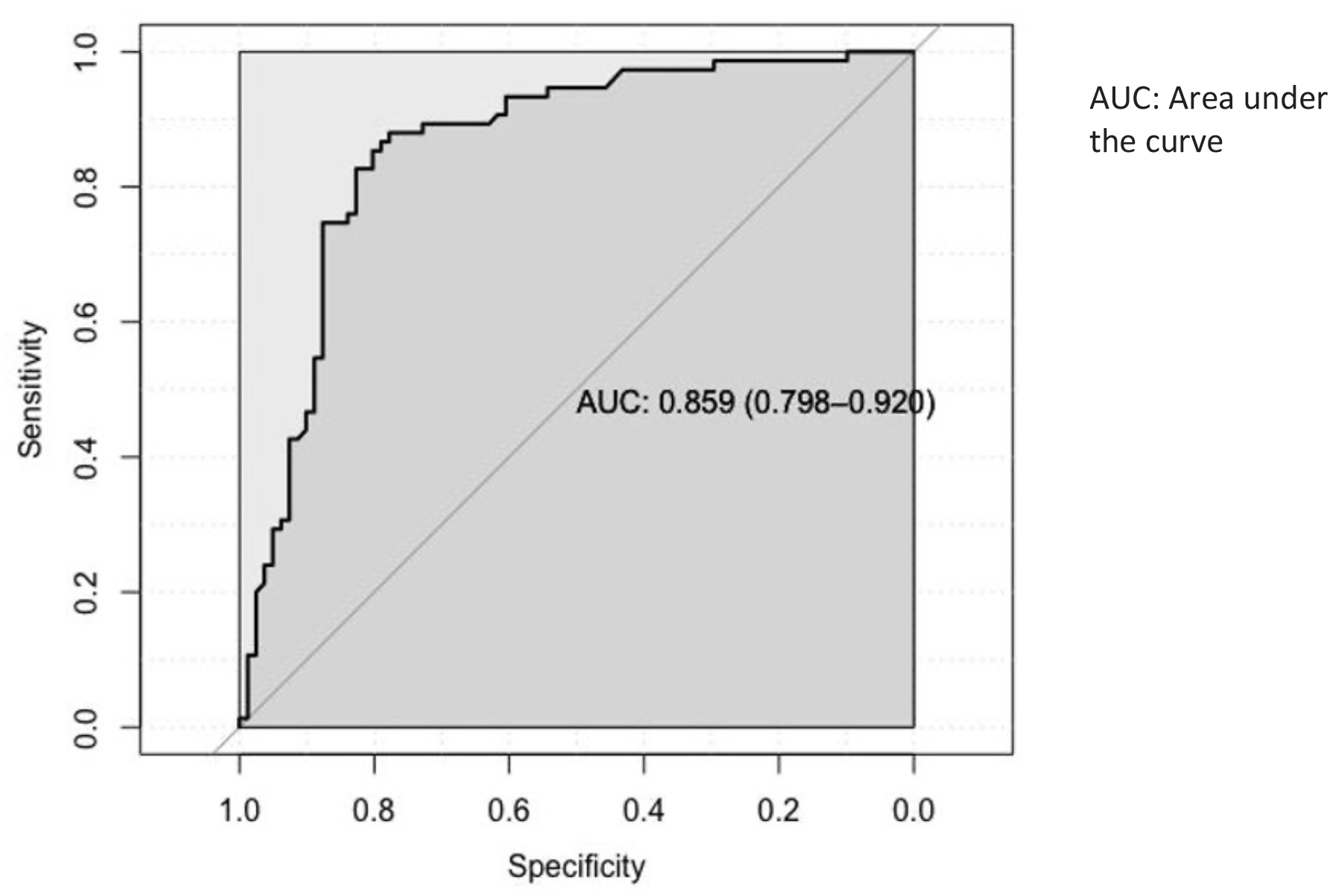
ROC curve for CAIOC-13 with OCD and Non-Clinical Control Group. The diagonal line indicates chance level.

### Factorial Validity

Using principal component analysis, the CAIOC-13 item scale’s dimensionality in the OCD group was examined. The number of factors to keep was decided using the scree test and the interpretability of the factor solution. A one-factor solution was indicated by the scree plot. Factor 1 accounted for 67 % (eigen value = 8.86) of the variance. Furthermore, after examining the component matrix of the factor analysis it became apparent that all items significantly loaded on only one factor, therefore it felt reasonable to select the unrotated one factor solution.

**Figure 2:**
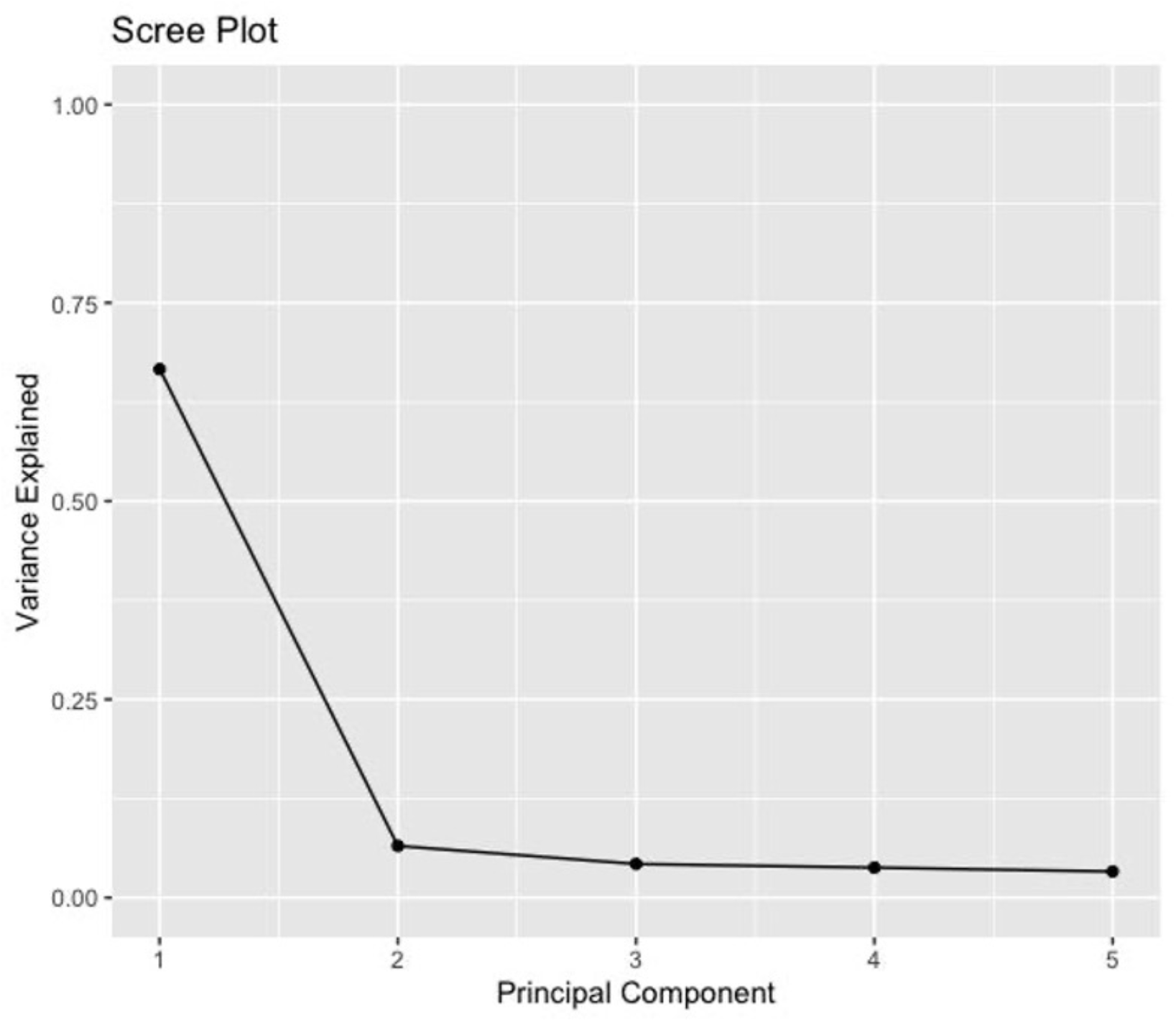
Scree Plot showing the variance explained by the components.

## DISCUSSION

This study was undertaken to establish the validity of the CAIOC-13 scale on an Indian sample of patients with OCD. The scale was self-rated, acceptable to patients and easy to apply. Importantly, it was sufficiently brief to be used in a clinical setting.

The study aimed to establish four types of validity for the CAIOC-13 scale: convergent, divergent, discriminant and factorial validity. The original validation research for the CAIOC-13 scale (15) was done on a sample of individuals drawn from the British population. There are some major differences between both the studies. The original study established concurrent validity by comparing CAIOC-13 with standard validated scales of OCD severity like the Clinical Global impressions Scale (CGI-S) and the Sheehan Disability Scale (SDS) which is self-rated scale of psychosocial disability. It also used secondary correlations with YBOCS and MADRS for concurrent validity. However, it did not compare with any other scale measuring cognitive deficits. Concurrent validity indicates whether a test that is designed to measure a particular construct correlated with other tests that assess the same or similar constructs (24). In the present study we used PDQ, which is also a self-rated tool of everyday cognitive dysfunction to establish concurrent validity. A correlation stronger than 0.5 is considered moderately strong in medical research (25). The present findings establish the concurrent validity of CAIOC-13 in Indian settings, through a significant positive correlation (r=0.526) with the PDQ. The PDQ assesses cognitive deficits in a variety of domains, including attention, retrospective memory, prospective memory, and planning and organisation. Even though both CAIOC-13 and PDQ are self-report measures of cognitive impairments in everyday life, PDQ was mainly developed to provide an assessment of domains of cognitive functioning that are specifically and frequently affected in Multiple Sclerosis (MS). The questionnaire has several items to measure prospective and retrospective memory deficits which may have less relevance for OCD. The CAIOC-13 is a more appropriate tool for the OCD sample given the domains assessed.

The original study did not establish divergent validity of CAIOC-13 with any other scale. Divergent validity is the requirement that the scale shows a weak correlation with dissimilar measures (24). Accordingly, the CAIOC-13 shows a low to moderate correlation (0.339) with the DAS-SF1 in our study. The DAS-SF1 assesses dysfunctional attitudes and cognitive biases in psychiatric populations (23). It was primarily developed for assessing dysfunctional cognitions in depression, subtyped into categories of Dependency, Self-control, and Achievement. A weak correlation with DAS-SF1 suggests that the CAIOC-13 distinctively measures the construct of subjectively perceived cognitive impairment rather than dysfunctional attitudes and cognitive biases.

The ROC analysis was used to establish the discriminant validity of the CAIOC-13 scale (26). The ROC graph was used to visualise and choose classifiers, that is, to assess the test’s capacity to appropriately categorise participants into clinically relevant subgroups based on performance (27). In the present study, findings from the ROC analyses suggest that the CAIOC-13 could be an effective measure for discriminating between patients with OCD and non-clinical controls. The optimal cut-off score in the OCD group was 28.5 which yielded high sensitivity and specificity, indicating that CAIOC-13 was able to identify cognitive impairments associated with OCD and not non-clinical controls. It is important to note that the optimal cut-off score reached in our sample is much lower than the threshold score of 38.5 reached in the initial study of the CAIOC-13 scale’s development (15). This suggests that the CAIOC-13 scale, when administered to a sample of Indian participants, can reliably discriminate between cognitive impairment caused by OCD and general functional dysfunction in the non-clinical controls at the cut-off score of 28.5. This difference may be attributed to the fact that only two groups have been used compared to the original study which had three groups: OCD, Depression and Healthy Controls. Another reason might have been the OCD severity in the present sample which ranged from moderate to severe cases (YBOCS range: 19 to 35) compared to the original sample which had several subclinical cases and severity range was much wider (between 5 to 36).

Another significant approach for determining the validity of a scale is factorial validity. It aids in examining the putative underlying structure of the scale and establishing its uni-dimensionality, where all the scale items cohere around a single latent dimension. The current study’s factor analysis results indicated that all CAIOC-13 items loaded significantly on a single factor of cognitive dysfunction, indicating that the scale is unidimensional and has a factor structure that is aggregated to produce a single construct.

CAIOC-13 is a valid tool for assessing functional cognitive impairment unique to OCD without undertaking time-, labour- and cost-intensive neuropsychological batteries. Evaluation of such functional cognitive impairment is important because cognitive dysfunction is prominent, persistent, and predictive of clinical outcomes in OCD (3,4,28–30). CAIOC-13 scale permits a brief and focussed record of these deficits as they impact functionality.

Overall, the findings from this study suggest that the CAIOC-13 is a successful and valid measure of the cognitive impact on functioning in patients with OCD in India. The results indicate that in addition to the clinical symptoms of obsessions and compulsions, cognitive dysfunction in the form of factors such as lassitude, slowness, indecisiveness, anxiety, procrastination, flexibility etc., also significantly contributes to clinical morbidity and impaired function.

### Limitation

The current study has several drawbacks. The study sample was limited to a primarily urban, English-speaking group, which may not accurately reflect the bulk of the Indian population and hence may impact the generalizability of the findings. Even though the sample size was calculated to be adequate for a medium effect size, a larger sample from a broader geographic area in India, with a wider age range may contribute to the psychometric properties of the scale.

The results can be strengthened by making comparisons with other groups of psychiatric diagnoses and demonstrating the scale’s level of specificity. Finally, because the scale was a self-report assessment and the data for the non-clinical controls was obtained online, there was a possibility of response bias, in which individuals replied erroneously or arbitrarily.

### Future directions

The CAIOC-13 aids in comprehensively analysing the day-to-day cognitive challenges experienced by patients suffering with OCD, which are not routinely examined by currently available tools. Over a variety of categories, most patients had moderate impairment. This is the first study administering the English-language CAIOC-13 scale in Indian adults, and therefore future studies are needed to assess the replicability of these findings. Future research will be needed to explicitly connect CAIOC-13 with neuropsychological laboratory tests that have previously shown to be impaired in OCD, such as assessments of response inhibition, set shifting and non-verbal memory. Larger sample studies may also potentially distinguish cognitive difficulties in distinct OCD symptom dimensions. The scale’s sensitivity to detect changes in functional impairment longitudinally and in response to intervention (such as cognitive training) are other important aspects to examine in future. Furthermore, in order to apply the scale effectively in the Indian community, it must be translated and empirically validated in other Indian languages.

## CONCLUSION

The current study has systematically established convergent, divergent, and discriminant validity and factorial structure of the CAIOC-13 on an Indian sample. Cognitive dysfunction in OCD may be marked, and contribute significantly to functional impairments, but may often be overlooked in busy clinical settings. The CAIOC-13 is thus of particular relevance to resource-constrained settings such as India.

## Data Availability

All data produced in the present study are available upon reasonable request to the authors

## ACKNOWLEDGEMENTS

We would like to thank all the participants in this study and the staff including the residents and the consultants of OCD clinic for referring patients for the study.

